# Glasses and risk of COVID-19 transmission - analysis of the Virus Watch Community Cohort study

**DOI:** 10.1101/2022.03.29.22272997

**Authors:** Annalan M D Navaratnam, Christopher O’Callaghan, Sarah Beale, Vincent Nguyen, Anna Aryee, Isobel Braithwaite, Thomas E Byrne, Wing Lam Erica Fong, Ellen Fragaszy, Cyril Geismar, Susan Hoskins, Jana Kovar, Parth Patel, Madhumita Shrotri, Sophie Weber, Alexei Yavlinsky, Robert W Aldridge, Andrew C Hayward, the Virus Watch Collaborative

## Abstract

**Background:** Respiratory viruses, including SARS-CoV-2, can infect the eyes or pass into the nose via the nasolacrimal duct. The importance of transmission via the eyes is unknown but might plausibly be reduced in those who wear glasses. Previous studies have mainly focussed on protective eyewear in healthcare settings.

**Methods:** Participants from the Virus Watch prospective community cohort study in England and Wales responded to a questionnaire on the use of glasses and contact lenses. This included frequency of use, purpose, and likelihood of wearing a mask with glasses. Infection was confirmed through data linkage with Second Generation Surveillance System (Pillar 1 and Pillar 2), weekly questionnaires to self-report positive polymerase chain reaction or lateral flow results, and, for a subgroup, monthly capillary blood testing for antibodies (nucleocapsid and spike). A multivariable logistic regression model, controlling for age, sex, income and occupation, was used to identify odds of infection depending on the frequency and purpose of using glasses or contact lenses.

**Findings:** 19,166 Virus Watch participants responded to the questionnaire, with 13,681 (71.3%, CI 70.7-72.0) reporting they wore glasses. A multivariable logistic regression model showed a 15% lower odds of infection for those who reported using glasses always for general use (OR 0.85, 95% 0.77-0.95, p = 0.002) compared to those who never wore glasses. The protective effect was reduced in those who said that wearing glasses interfered with mask wearing. No protective effect was seen for contact lens wearers.

**Interpretation:** People who wear glasses have a moderate reduction in risk of COVID-19 infection highlighting the importance of the eye as a route of infection. Eye protection may make a valuable contribution to the reduction of transmission in community and healthcare settings.

**Funding:** The research costs for the study have been supported by the MRC Grant Ref: MC_PC 19070 awarded to UCL on 30 March 2020 and MRC Grant Ref: MR/V028375/1 awarded on 17 August 2020. The study also received $15,000 of Facebook advertising credit to support a pilot social media recruitment campaign on 18th August 2020. The study also received funding from the UK Government Department of Health and Social Care’s Vaccine Evaluation Programme to provide monthly Thriva antibody tests to adult participants. This study was supported by the Wellcome Trust through a Wellcome Clinical Research Career Development Fellowship to RA [206602]. Funding from the HSE Protect study, GOSH Children’s Charity and the Great Ormond Street Hospital BRC supported the involvement of CO in the project.

**Research in context:** *Evidence before the study:* Despite the risk of SARS-CoV-2 transmission via the eyes, very few countries have advocated eye protection to reduce transmission amongst the public and, except when providing close care for those known or suspected to be infected, is variable and based on case-by-case assessment of exposure risk. The mechanism, but not the extent, of the transmission route through the eyes is well described in the literature, with several studies reporting detection of SARS-CoV-2 RNA in the tear film, conjunctiva and conjunctival sac. There have been a small number of hospital based observational studies suggesting that eye protection may help prevent COVID-19 infection. A literature search was carried out on 23rd February 2022 across Medline and Embase using the search terms ‘eyewear’, ‘glasses’, ‘SARS-CoV-2’, ‘COVID-19’, ‘SARS’, ‘transmission’ and ‘infectivity’, providing 105 manuscripts. Of these, only eight investigated the risk of infection associated with eye protection, all in hospital settings or followed a cohort of healthcare workers. Among the studies was a systematic review that identified 5 observational studies from 898 articles that were screened. The cohort study with the largest sample size, 345 healthcare professionals, demonstrated a relative risk of 10.25 (95% CI 1.28–82.39; P = 0.009) for infection when not using eye protection. No studies of the potential protective effect of glasses wearing, for visual correction, in community settings were identified.

*Added value of this study:* The Virus Watch study is a prospective community household study across England and Wales. 19,166 participants responded to the monthly questionnaire on glasses and contact lens use, assessing reported frequency, the purpose of use and how likely they were to wear a mask with glasses. Infections were identified in data linked to the Second Generation Surveillance System (Pillar 1 and Pillar 2 testing), weekly surveys seeking self-reports of polymerase chain reaction or lateral flow device results and, in a subset of 11,701, self-collected capillary blood testing for antibodies (nucleocapsid and spike - nucleocapsid antibodies were taken as evidence of prior infection as these are unaffected by vaccination). Our multivariable logistic regression model, controlling for age, sex, household income and occupation, demonstrated 15% lower odds of infection for those who reported always using glasses for general use compared to those who never wear glasses. The protective effect was not observed in those who strongly agreed with the statement, ‘I am less likely to wear a face covering when I have my glasses on because my glasses steam up’. Counterfactual analysis of contact lenses did not suggest a protective effect regardless of frequency of use.

*Implications of all the available evidence:* The findings of this study demonstrate a moderate reduction in risk of SARS-CoV-2 infection in those who always wear glasses compared to never. Unlike other studies, our results are representative of a community setting, adjust for potential confounders and provide a counterfactual analysis with contact lenses. This extends the current evidence to community settings and validates proposed biological mechanisms of eye protection reducing the risk of SARS-CoV-2 transmission.

## Background

Respiratory viruses infect individuals via the nose, mouth and eyes, through contact with surfaces touched by the individual or via small and larger (i.e. droplet) aerosol particles.^1^ Recommendations for the protection of the general public in most countries include social distancing, handwashing and face mask use but not eye protection. In the UK eye protection (including full face visors or goggles) is recommended in healthcare settings if blood or body fluid contamination to the eyes or face is anticipated or likely. Also if caring for patients with a suspected or confirmed infection spread by the droplet or airborne route as deemed necessary by a risk assessment, or during aerosol generating procedures.^2^ Regular corrective glasses are not considered as eye protection.

The eyes present two routes for SARS-CoV-2 infection, the first through infection of conjunctival cells that contain ACE2 receptors. Several studies have detected SARS-CoV-2 RNA in the tear film, conjunctiva and conjunctival sac with between 1-12% of patients with COVID-19 reported to have ocular manifestations.^3–6^ The second infection route is via the nasolacrimal duct, which is known to transport pathogens to the nose within minutes and onward to the nasopharynx.^7^ Supporting the eye as a route of SARS-CoV-2 infection, conjunctival inoculation of the virus in macaques leads to interstitial pneumonia in macaques.^8^ A small number of hospital based observational studies suggest that eye protection may help prevent COVID-19.^9^ Amongst these, an observational study of 276 COVID-19 patients admitted to a hospital found the proportion of spectacle wearers was lower than the general population.^10^

Based on the biological mechanisms and studies in healthcare we hypothesised that glasses wearing in community settings would reduce the risk of COVID-19. Glasses may provide a barrier to prevent exposure to infectious aerosol particles, particularly the ballistic component of larger particles, and may also reduce contaminated fingers touching the eyes. We do not expect to see this same protective effect in a counterfactual contact lens analysis. We therefore developed a survey on glasses and contact lenses within the Virus Watch cohort to test these hypotheses. The aim of this study was to test the hypothesis that wearing glasses is associated with a lower risk of COVID-19.

## Methods

The Virus Watch study is a household community cohort of acute respiratory infections in England and Wales that started recruitment in June 2020.^11^ As of 2nd February 2022, 58,670 participants were recruited using a range of methods including post, social media and SMS messages and letters from their General Practice. Participants provided a wide range of information on registration including age, sex, occupation and household information (e.g. household income, postcode). In the December 2022 monthly questionnaire, 31,749 participants were asked whether they used glasses or contact lenses, and if so, how frequently they used them generally, for reading or for carrying out a specific task. They were also asked about the level of agreement with the following statement: ‘I am less likely to wear a face covering when I have my glasses on because my glasses steam up’.

The covariates considered in this analysis were age, sex, income and occupation. Income was defined by the combined household income divided by the number of adults in that household. This was then put into categories ranging from £0-9,999 to £80,000+, with intervals of £10,000. Self-reported occupation was grouped using the Office for National Statistics’ Standard Occupational Classification Hierarchy.^12^ ‘Skilled trades occupations’ and ‘Process, plant and machine operatives’ were grouped into ‘manual’, with all other occupations grouped as ‘non-manual’. If occupation was not available, ‘not reported’ was recorded.

There were multiple ways in which to identify the first SARS-CoV-2 infection among participants of this study. Infection was identified based on the first positive result from the following sources:

1. Data linked to the Second Generation Surveillance System (SGSS), which contains SARS-CoV-2 test results using data from hospitalisations (Pillar 1) and community testing (Pillar 2). Linkage was conducted by NHS Digital with the linkage variables being sent in March 2021.The linkage period for SGSS Pillar 1 encompassed data from March 2020 until August 2021 and from June 2020 until November 2021 for Pillar 2.
2. Self-reported positive polymerase chain reaction (PCR) or lateral flow device (LFD) swabs for SARS-CoV-2 infection as part of the Virus Watch weekly survey.
3. Monthly self-collected capillary blood samples (400-600μL) in a subsample of 11,701 participants, which were tested in United Kingdom Accreditation Service (UKAS)-accredited laboratories. Serological testing using Roche’s Elecsys Anti-SARS-CoV-2 electrochemiluminescence assays targeting total immunoglobulin (Ig) to the Nucleocapsid (N) protein, or to the receptor binding domain in the S1 subunit of the Spike protein (S) (Roche Diagnostics, Basel, Switzerland). At the manufacturer-recommended seropositivity thresholds (≥1.0 cut-off index [COI] for N and ≥0.8 units per millilitre [U/ml] for S). A positive result was defined based on positivity to the N protein.
4. Clinical-collected venous blood samples tested for the S protein. In-clinic serology was conducted twice per participant between September 2020-January 2021 (Autumn round n = 3050) and April 2021-July 2021 (Spring round n = 2775)) (see study protocol for details).^11^ Positivity was defined as evidence of S-positivity in absence of receiving any COVID-19 vaccination prior to the serological test.

We used sliding date window matching (14 day window) to identify positive tests recorded by both Virus Watch and linkage to UK national records; where both were available, the linkage date was used. Where both swab and serological positives were recorded, we used the PCR/LFT date, unless the serological positive occurred first. Reinfections were not included.

### Outcomes

The primary outcome is the risk of infection depending on self-reported use of glasses, grouped into frequency of use. Frequency of use in the questionnaire was reported as ‘Never’, ‘Rarely’, ‘Sometimes’, ‘Most times’ and ‘Always’. ‘Rarely’, ‘Sometimes’ and ‘Most times’ were then grouped into ‘Sometimes’.

Secondary outcomes were risk of infection depending on the use of mask and glasses at the same time as well as the frequency of use of contact lens (for counterfactual analysis).

### Analysis

All individuals who responded to the December 2022 monthly survey were included in the analysis. Any individuals with a positive test, by any of the methods mentioned previously, were considered to have had an infection. It was assumed that if participants did not have a positive test, they would not have had prior SARS-CoV-2 infection. The first date of infection was used in this analysis and subsequent infections were excluded. Proportions of positive individuals were calculated with 95% confidence intervals. Multivariable logistic regression models were created, which included age (as a continuous variable), sex, household income per adult in the household and occupation, grouped into manual, non-manual and not reported. For comparison of binary variables, the Chi-Square test was used. All analyses were carried out with R studio (R 4.0.5.) using packages; ‘tidyverse’, ‘ggplot2’ and ‘rstatix’.

## Results

Of 58,670 participants in the Virus Watch cohort, 31,749 were invited to answer the monthly survey on glasses and contact lens use. There were 19,166 respondents to this questionnaire. The median age was 63 years old (IQR 52-70) and 10,470 of participants were female (54.6%, 95% CI 53.9-55.3%). A total of 13,681 participants (71.3%, CI 70.7-72.0) reported wearing glasses. Of 19,166, 19.6% (3,757, 95% CI 19.0-20.2) had evidence of prior COVID-19 infection. There was also no difference between sex, with 19.6% (8,255, 95% CI 18.7-20.5) of males and 19.9% females (10,470, 95%CI 19.1-20.6) having evidence of a previous infection.

Amongst those who never wore glasses for general use, 22.99% (95% CI 22.01-23.97%) were infected vs 15.63% (95% CI 14.76-16.5%) for those who always wore glasses for general use. (Table 1). The multivariable logistic regression model, which included age, sex, household income per adult and occupation, showed a 15% lower odds of infection for those who reported using glasses always for general use (OR 0.85, 95% 0.77-0.94, p = 0.002) compared to non-wearers (Figure 1). A similar pattern was seen with using glasses always for reading and other specific tasks. There was no evidence of protection from wearing glasses only sometimes and no evidence of protection from any frequency of wearing contact lenses (Figure 1).

**Table 1:** Summary table of proportion of individuals with prior SARS-CoV-2 infection grouped by type and frequency of use with 95% confidence intervals. Missing refers to missing data.

| **Eyewear** | **Usage** | **Frequency** | **Total Responses** | **Positives n (%)** | **95%CI** |
| --- | --- | --- | --- | --- | --- |
| Glasses | General use | Never | 7047 | 1620 (22.99) | 22.01, 23.97 |
|  |  | Sometimes | 4959 | 1002 (20.21) | 19.09, 21.32 |
|  |  | Always | 6687 | 1045 (15.63) | 14.76, 16.5 |
|  |  | Missing | 473 | 90 (19.03) | 15.49, 22.56 |
|  | Other specific | Never | 7077 | 1640 (23.17) | 22.19, 24.16 |
|  |  | Sometimes | 3383 | 711 (21.02) | 19.64, 22.39 |
|  |  | Always | 7405 | 1172 (15.83) | 15, 16.66 |
|  |  | Missing | 1301 | 234 (17.99) | 15.9, 20.07 |
|  | Reading | Never | 5934 | 1401 (23.61) | 22.53, 24.69 |
|  |  | Sometimes | 4695 | 946 (20.15) | 19, 21.3 |
|  |  | Always | 7948 | 1299 (16.34) | 15.53, 17.16 |
|  |  | Missing | 589 | 111 (18.85) | 15.69, 22 |
| Contact Lenses | General use | Never | 15718 | 3074 (19.56) | 18.94, 20.18 |
|  |  | Sometimes | 1333 | 303 (22.73) | 20.48, 24.98 |
|  |  | Always | 1131 | 222 (19.63) | 17.31, 21.94 |
|  |  | Missing | 984 | 158 (16.06) | 13.76, 18.35 |
|  | Other specific | Never | 15291 | 3007 (19.67) | 19.04, 20.3 |
|  |  | Sometimes | 1162 | 257 (22.12) | 19.73, 24.5 |
|  |  | Always | 1086 | 215 (19.8) | 17.43, 22.17 |
|  |  | Missing | 1627 | 278 (17.09) | 15.26, 18.92 |
|  | Reading | Never | 15422 | 3036 (19.69) | 19.06, 20.31 |
|  |  | Sometimes | 1168 | 256 (21.92) | 19.55, 24.29 |
|  |  | Always | 1096 | 214 (19.53) | 17.18, 21.87 |
|  |  | Missing | 1480 | 251 (16.96) | 15.05, 18.87 |

**Figure 1.**
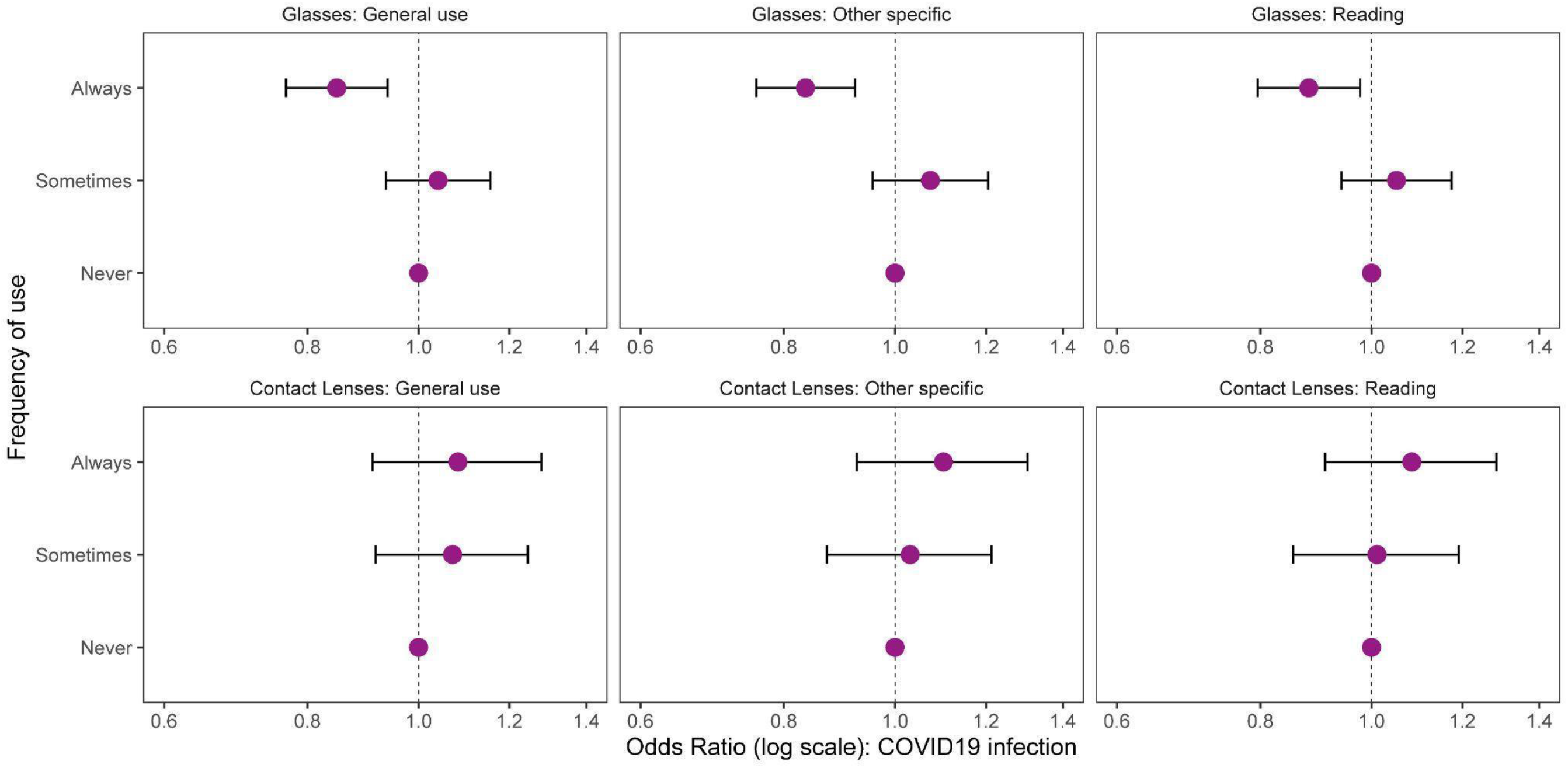
Adjusted odds ratios and 95% confidence intervals showing association of glasses and contact use with COVID-19 infection. Adjusted for age, sex, income and occupation.

When glasses wearers were asked if they agreed with the statement ‘I am less likely to wear a face covering when I have my glasses on because my glasses steam up’ there was evidence that when glasses interfered with mask use there was a reduction in the protective effect (Table 2 and Figure 2).

**Table 2:** Summary table of proportion of individuals with prior SARS-CoV-2 infection grouped by level of agreement with ‘I am less likely to wear a face covering when I have my glasses on because my glasses steam up’’ with 95% confidence intervals.

| Response | Total Responses | Positives (%) | 95%CI |
| --- | --- | --- | --- |
| Strongly Disagree | 6257 | 958 (15.31) | 14.42, 16.2 |
| Disagree | 3336 | 608 (18.23) | 16.92, 19.54 |
| Neither | 1574 | 311 (19.76) | 17.79, 21.73 |
| Agree | 1774 | 387 (21.82) | 19.89, 23.74 |
| Strongly Agree | 534 | 134 (25.09) | 21.42, 28.77 |
| Missing data | 5691 | 1359 (23.88) | 22.77, 24.99 |

**Figure 2:**
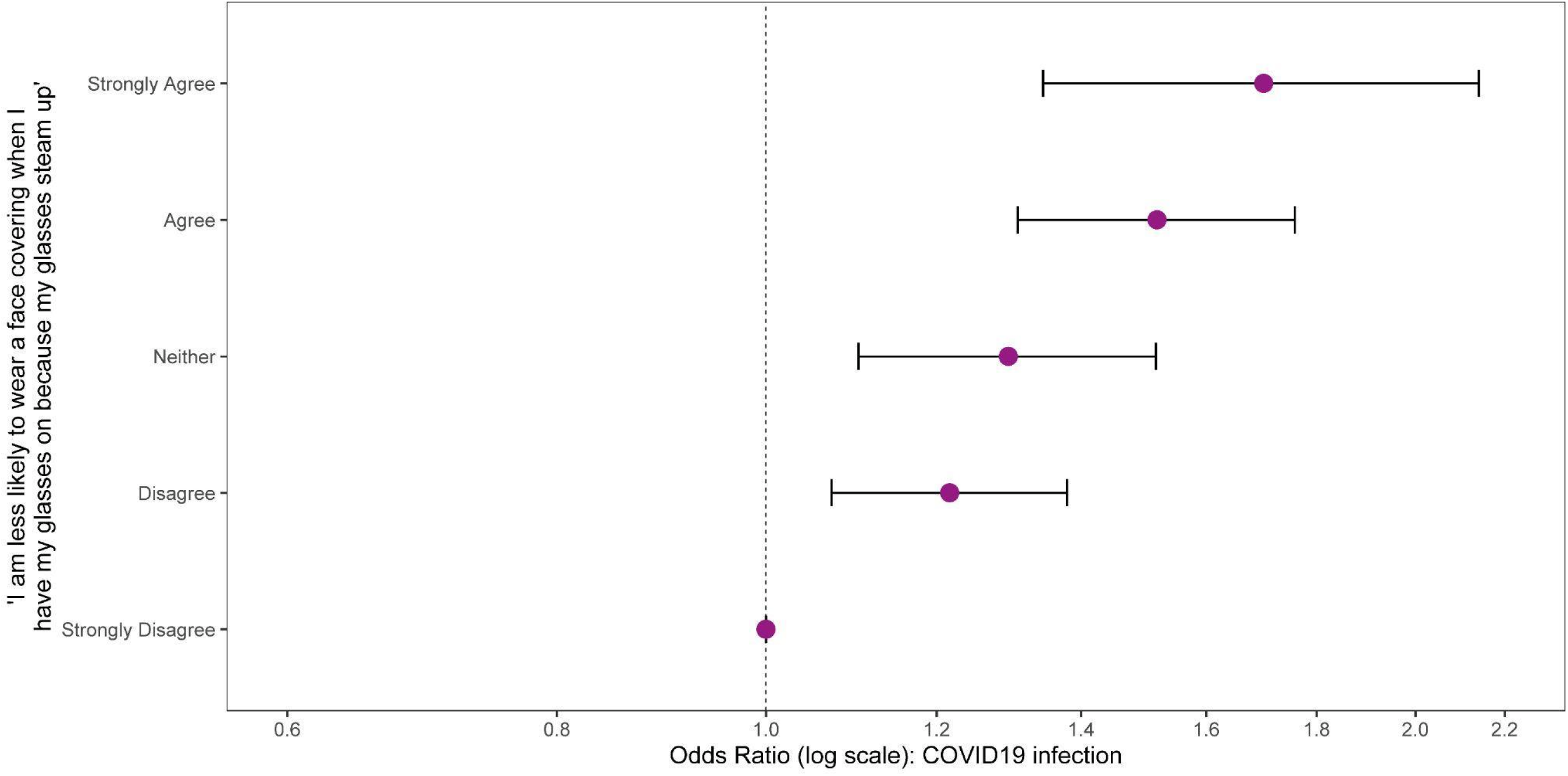
Adjusted odds ratios and 95% Confidence intervals comparing glasses users according to their agreement with the question “I am less likely to wear a face covering when I have my glasses on because my glasses steam up”. Adjusted for age, sex, income and occupation.

## Discussion

Our results show a significant reduction in the risk of COVID-19 infection among those who always wear glasses. This demonstrates the importance of the eyes as a portal for infection and suggests that strategies to broaden the use of eye protection could help to prevent transmission and contribute to infection control. The counterfactual analysis of contact lenses showed no protective effect, strongly suggesting a causal relationship to glasses wearing and reduced risk of SARS-CoV-2 infection. The findings also highlight that many individuals reduce mask wearing with glasses, because of glasses steaming up, which is associated with a reduced protective effect. This suggests the need for mask design and usage to prevent steaming up during use as well as adds to growing evidence of face coverings’ protective effect.

There are a number of plausible mechanisms by which wearing glasses could contribute to prevention of COVID-19 infection. Healthy individuals involuntarily touch their eyes around three times per hour and wearing spectacles may reduce the number of times SARS-CoV-2 contaminated fingers touch the eyes.^13^ It is likely that spectacles present a barrier to the direct impaction of viruses on the eyes. Eye deposition of SARS-CoV-2 may also occur directly from the impaction of the ballistic component of aerosol particles, particularly larger particles (i.e. droplets) produced by coughing and sneezing. Air currents may also direct virus containing aerosolised particles towards the eye, enhancing deposition, and Brownian motion of aerosolised particles may also result in deposition on the ocular surface.

We hypothesise that even greater protection against COVID-19 may be afforded by eye protection that wraps around the eyes or seals the eyes from the environment. Face shields are frequently used in hospitals and by the public in some countries. Like glasses, they may reduce infection risk to the ballistic component of droplets, but they do not offer full protection against small aerosol particles, as illustrated by experimental studies.^14^ Protection afforded by eye protection is also likely to be seen for other respiratory viruses, such as influenza and respiratory syncytial virus (RSV), which remain infectious in exhaled aerosol particles. ^15,16^

The counterfactual absence of a protective effect in contact lens wearers also helps to strengthen the findings. The absence of protective effect with contact lens aligns with the biological mechanisms of SARS-CoV-2 infection through the eyes. Hands are a vector of transmission, and using contact lens is associated with increase contamination from fingertips due to application, removal, and adjustment (because of dry eyes and irritation) of lenses.^17–20^ Furthermore, as contact lenses only cover the cornea of the eye, so there is no protection from the two routes of infection; the conjunctiva and nasolacrimal ducts.

Strengths of this work include the prospective approach, large numbers of participants, multiple approaches to capture SARS-CoV-2 infections and unlike previous studies, the ability to control for potential confounders.^9^ We considered the need for visual correction is strongly influenced by age but not by other variables that may impact on risk of infection. However, choice of contact lenses versus glasses may be affected by social factors and occupation and poor visual acuity may prevent people from working in some occupations. It was therefore, important to be able to control for these variables. As our questions are specific to eye glasses, not inclusive of face shields, we have overcome concerns of whether the reduced transmission is through reduced inhalation or protecting eyes.^9^ An assumption in our analysis however is that the reported frequency of use was reflective of the entire study period. We also note that the benefit of glasses wearing may be greater in those with the most exposure (e.g. those unable to work from home or working in health care facilities), but we did not have sufficient power to explore these hypotheses.

Although community based, the findings of this study show the potential of eye protection to reduce infection risk and may be particularly important in high exposure settings such as healthcare. Eye protection has been reported to be the most frequently missed item of PPE among healthcare workers, emphasising the importance of providing evidence on its benefits.^21^ Our work adds to existing observational studies extending the evidence of the protective effects beyond healthcare workers and into the community settings.

## Conclusion

Extending the use of protective eyewear should be considered as part of broader strategies to prevent community transmission of infection and may be valuable to consider in the event of future pandemics and in high exposure occupations including healthcare.

## Data Availability

All data produced in the present study are available upon reasonable request to the authors.

## Ethics

This study has been approved by the Hampstead NHS Health Research Authority Ethics Committee. Ethics approval number - 20/HRA/2320.

## Data availability

We aim to share aggregate data from this project on our website and via a “Findings so far” section on our website - https://ucl-virus-watch.net/. We will also be sharing individual record level data on a research data sharing service such as the Office of National Statistics Secure Research Service. In sharing the data we will work within the principles set out in the UKRI Guidance on best practice in the management of research data. Access to use of the data whilst research is being conducted will be managed by the Chief Investigators (ACH and RWA) in accordance with the principles set out in the UKRI guidance on best practice in the management of research data. We will put analysis code on publicly available repositories to enable their reuse.

